# Part 2: Predicting heterogeneity of treatment effects to transcranial direct current stimulation for knee osteoarthritis pain and symptoms

**DOI:** 10.1101/2025.06.09.25329238

**Authors:** Chiyoung Lee, Heewon Kim, Seoyoung Kim, Yeri Kim, Xiaoxiao Sun, Chen X. Chen, Juyoung Park, Christine Pellegrini, David O. Garcia, Nan-kuei Chen, C. Kent Kwoh, Hyochol Ahn

**Author notes:** **Correspondence: Chiyoung Lee, PhD, RN** Assistant Professor, The University of Arizona College of Nursing 1305 N Martin Avenue, Tucson, AZ 85721-0203.

## Abstract

**Background:** Assessing the heterogeneity of treatment effects (HTE) is a fundamental aspect of precision medicine, which aims to predict the most optimal treatments based on participant-specific characteristics. This study seeks to identify key predictors of the HTE of transcranial direct current stimulation (tDCS) in individuals with symptomatic knee osteoarthritis (KOA) using machine-learning approaches.

**Methods:** We performed a secondary analysis of a randomized clinical trial involving 60 participants with symptomatic KOA. These participants underwent 15 daily sessions of 2-mA active tDCS (each session lasting 20 minutes) over a period of three weeks. Initially, we applied group-based trajectory modeling to classify participants into distinct subgroups based on longitudinal KOA pain and symptom patterns from baseline to three months post-intervention to examine differential responses to tDCS. A multi-layer perceptron classifier was then trained to predict the trajectory subgroups using demographic, clinical, and quantitative sensory testing data collected during baseline visits. Feature selection methods, including f-regression, r-regression, and SHapley Additive Explanations (SHAP), were employed to identify the influential features. Additionally, SHAP was used to analyze the correlation and impact of each feature on classification.

**Results:** Participants exhibited distinct response patterns to tDCS: high responders (individuals with low initial symptoms showing significant improvement, *n* = 28) and low responders (individuals with high initial symptoms showing minimal improvement, *n* = 32) to tDCS. The influential features included conditioned pain modulation (CPM), cold pain intensity, pressure pain thresholds (PPTh) at the medial knee and trapezius, and pain catastrophizing. SHAP analysis revealed that pain catastrophizing was the most influential feature. Additionally, lower CPM, higher cold pain intensity, lower PPTh, and greater pain catastrophizing were associated with a higher likelihood of being classified as low responders.

**Conclusion:** Our results contribute to the existing literature, suggesting that factors such as pain catastrophizing, peripheral and central pain sensitization, and individuals’ endogenous pain-inhibitory capacity should be carefully considered in future tDCS trials.

## 1. Introduction

Osteoarthritis is a leading cause of pain and disability in older adults, with knee osteoarthritis (KOA) being the most prevalent form, followed by hand and hip osteoarthritis.^1,2^ The chronic pain associated with KOA not only affects individuals but also places a significant strain on healthcare systems in terms of resources and costs.^3–5^ Given its high prevalence and substantial personal and societal consequences, there is a pressing need for effective and accessible treatment options.

As evidence continues to support the role of centrally mediated mechanisms in chronic KOA pain, interventions targeting pain-related brain regions,^6,7^ particularly transcranial direct current stimulation (tDCS), have gained recognition.^8^ Clinical trials, including findings from our research team, have shown promising results in the efficacy of tDCS in managing chronic KOA pain and symptoms.^9–13^ The primary motor cortex (M1), a key region involved in motor control and central pain processing, is the most commonly targeted brain area in tDCS.^14^ By delivering low doses of direct electrical current to the brain through electrodes on the scalp, tDCS induces changes in cortical neuronal activity.^8^

In the realm of tDCS research for KOA, the average treatment effect has traditionally been a guiding factor in evidence-based clinical decisions. When applying estimated average treatment effects, we implicitly assume that these effects are universally applicable to all individuals. However, it has become increasingly evident that some patients in active treatment groups may not respond positively or even experience deterioration, despite the average treatment effect indicating beneficial effects. Focusing solely on ATE may result in these patients missing out on potential treatment benefits. In light of the shift toward personalized medicine, the importance of assessing the heterogeneity of treatment effects (HTE) has gained widespread recognition. HTE involves the quantification of how patients respond differently to treatments.^15^ Identifying HTE is critical to ensure that treatment is administered optimally.^16^

Predictive modeling approaches to HTE analyses offer individualized predictions of treatment benefits by considering multiple relevant patient characteristics simultaneously, which are foundational to personalized medicine.^17^ By leveraging patient data, researchers can uncover patterns and predictors of treatment efficacy, thereby enabling more tailored and personalized interventions. Recently, machine-learning (ML) models have been increasingly utilized to predict varying responses to medical treatment.^18^ Due to the need to integrate variables from different fields to build predictive models, ML offers a data-driven approach that can incorporate a large number of variables into a single model; compared to traditional methods, ML can handle multicollinearity between variables without making assumptions about these associations and provide more precise predictions at the individual level in the era of personalized medicine.^18^

This study aimed to identify key predictors of the HTE of tDCS in individuals with symptomatic KOA. In our previous research (blind for review), the presence of HTE in this clinical population was demonstrated by identifying differential responses to tDCS, leading to the population’s classification into high responders (exhibiting low initial symptoms with significant improvement) and low responders (showing high initial symptoms with minimal improvement). The current study employed ML models trained on baseline demographic, clinical, and quantitative sensory testing (QST) data to identify the influential features for predicting the previously identified response trajectories.

## 2. Methods

### 2.1. Design

The current study involved a secondary analysis of a double-blind, randomized, sham-controlled, phase II pilot clinical trial with a parallel-group design. The original trial, registered on ClinicalTrials.gov (NCT04016272), consisted of a three-week home-based tDCS program monitored in real time via secure videoconferencing. Upon providing informed written consent, a total of 120 participants were randomly allocated into two groups: active tDCS and sham tDCS, with 60 participants assigned to each group (**Figure 1**). This study used data from the active tDCS group.

**Figure 1.**
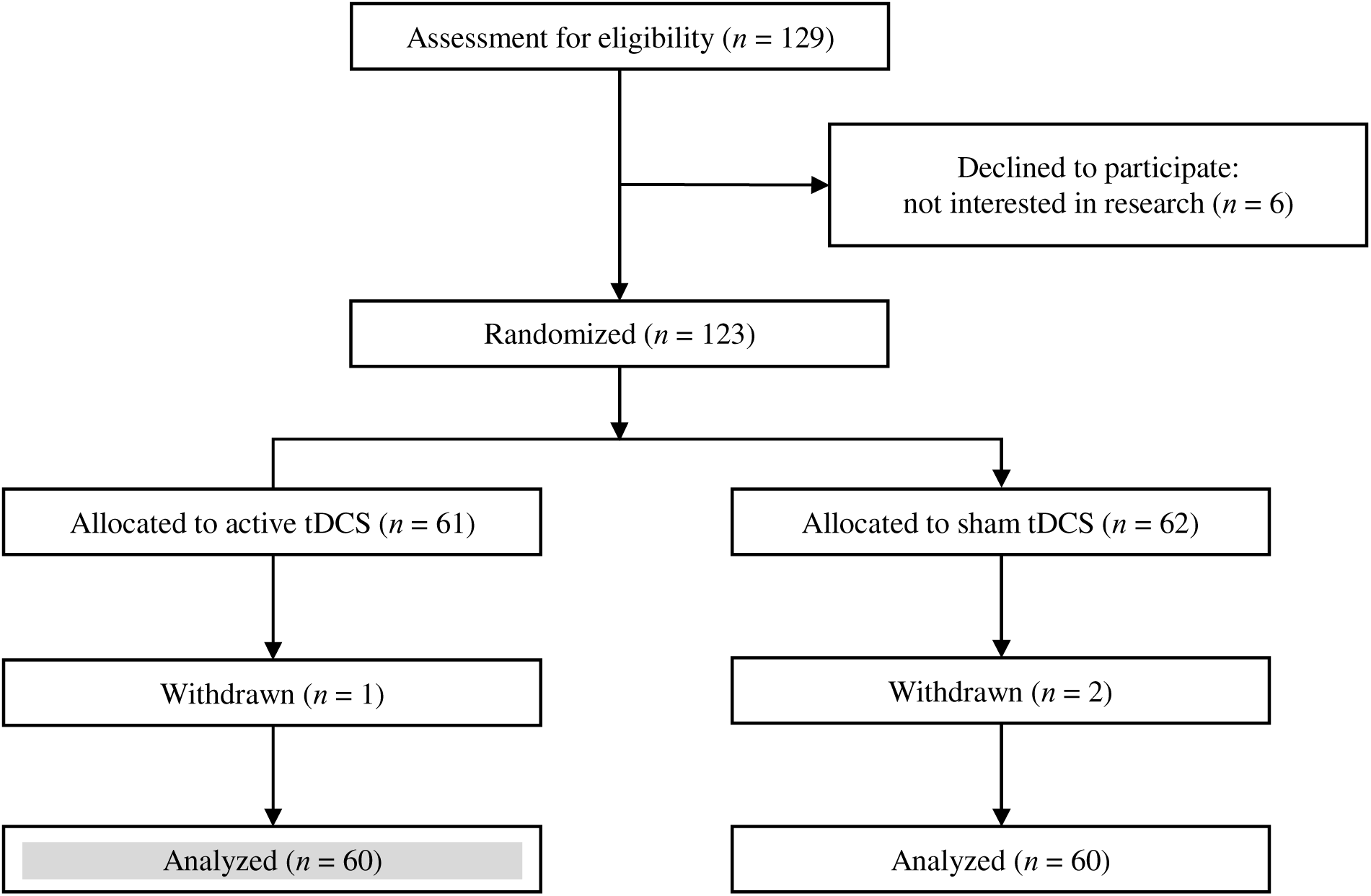
Participant flow diagram (*n* =120) *Note.* “Declined” meant that participants came to baseline visit (signed informed consent) but did not start any intervention. For this study, among 6 declined subjects, 4 subjects signed informed consent at the baseline but declined to study participation; 2 subjects did not show up at the baseline. <u>This study used</u> <u>data from the active tDCS group.</u>

### 2.2. Participants

Participants were recruited from Southeast Texas through advertisements at local institutions, including UTHealth, and flyers distributed in nearby communities. Additionally, potential participants were identified and directly approached for recruitment at the UT Physicians Orthopedics Clinic.

Participants aged 50 to 85 were eligible if they (1) have unilateral or bilateral symptomatic knee OA based on American College of Rheumatology Clinical criteria (i.e., knee pain, plus at least 3 of the following 6 signs/symptoms: age ≥ 50 years, morning stiffness < 30 min, crepitus, bony tenderness, bony enlargement, no palpable warmth),^19^ (2) have had KOA pain in the past three months with an average of at least 30 on a 0-100 Numerical Rating Scale (NRS) for pain,^20^ (3) can speak and read English, and (4) have no plans to alter their pain medication regimen during the study. Participants were excluded if they have any concurrent medical conditions that could confound the interpretation of outcome measures, pose a safety risk for any of the assessments or interventions, or hinder the completion of the protocol. Specific exclusion criteria are: (1) history of brain surgery, brain tumor, seizure, stroke, or intracranial metal implantation, (2) systemic rheumatic disorders, including rheumatoid arthritis, systemic lupus erythematosus, and fibromyalgia, (3) alcohol/substance abuse, (4) diminished cognitive function that would interfere with understanding study procedures (i.e., Mini-Mental Status Exam score ≤ 23), (5) pregnancy or lactation, (6) prosthetic knee replacement or non-arthroscopic surgery to the affected knee, and (9) hospitalization within the preceding year for psychiatric illness.

### 2.3. Intervention: Active tDCS

For chronic pain treatment, tDCS has been recognized to be successful when delivered with the anode electrode placed over the primary motor cortex (M1) and with the cathode electrode placed over the supraorbital region (SO).^21–23^ International experts in clinical neurophysiology recommend 20-minute M1-SO stimulation using 2 mA electrical current intensity for possible efficacy among populations with chronic pain.^23^ For active stimulation, tDCS with a constant current intensity of 2 mA was applied for 20 minutes per session via the Soterix 1×1 tDCS mini-CT Stimulator device (Soterix Medical Inc., NY; 6.5 inches long, 3 inches wide, 0.7 inches thick) with headgear and 5×7 cm saline-soaked surface sponge electrodes. The sponge electrodes snap into the custom headgear, which is secured to the participant’s head for simple and fail-safe electrode preparation. This single-position headgear with clearly labeled sponge markers eliminates room for user error and helps maintain the placement of the montage. Participants can only administer a stimulation session via the Soterix 1×1 tDCS mini-CT Stimulator device after being provided a single-use unlock code by the research staff once proper placement and contact are achieved (only the on/off button is adjustable by the study participants; they cannot adjust the device settings). After the participant entered the unlock code, the screen on the device shows a timer that counts down the minutes until the end of the session. At 20 minutes, the device turns off automatically. The participant was instructed to remove the headset, discard the sponges, and safely store all materials for the next session.

### 2.4. Measurements

#### 2.4.1. KOA Pain and Symptoms

KOA symptom assessments were performed at specific time points, including baseline, after five intervention sessions (day 5), after an additional five sessions (day 10), and upon completion of the final set of five sessions (day 15). Follow-up evaluations were conducted at one month (day 45), two months (day 75), and three months (day 105) post-intervention.

##### Numeric Rating Scale (NRS)

Clinical pain intensity was measured by asking participants to rate their average knee pain over the past 24 hours via a NRS from 0 (no pain) to 100 (worst pain imaginable). This type of scale is recognized for its reliability, validity and sensitivity in various populations and settings, ^24–26^ including in adults with KOA.^11,27,28^

##### The Western Ontario and McMaster Universities Osteoarthritis Index (WOMAC)

We measured key joint symptoms using the Western Ontario and McMaster Universities Osteoarthritis Index (WOMAC),^29^ which consists of 3 subscales relating to pain during activities (range 0–20), stiffness during the day (range 0–8), and impairments of physical function (range 0–68), with higher scores indicating worse pain, stiffness, and impairments of physical function. Total scores range from 0 to 96, with higher scores indicating worse OA pain-related symptoms. WOMAC is a disease-specific, valid, and reliable instrument.

#### 2.4.2. Baseline Characteristics

Demographic characteristics, including age (continuous), gender (male *vs.* female), body mass index (BMI; kg/m²), race (white *vs*. non-white), education (high school or less *vs.* college or more), and marital status (married/partnered *vs.* non-married/unpartnered), as well as clinical characteristics, such as the index knee (the most affected knee), Kellgren-Lawrence score, and the average duration of osteoarthritis (months), were collected. Pain catastrophizing was measured via the Pain Catastrophizing Scale (PCS, ranging from 0 to 52), which consists of 13 items.^30^ It has excellent reliability with Cronbach’s alpha coefficient of ≥ 0.9 and is a well-validated measure.^31^

The QST methods involve the administration of various stimulus modalities (e.g., thermal and mechanical) and the evaluation of various perceptual endpoints (e.g., threshold, tolerance, and suprathreshold scaling).^32^ Furthermore, approaches for assessing pain modulatory function, including both inhibition and facilitation, are increasingly used.^32^ Generally, a multimodal QST protocol is recommended for a more comprehensive characterization of pain processing in clinical populations. In this study, the tests included heat pain threshold (HPTh), heat pain tolerance (HPTo), pressure pain threshold (PPTh), punctate mechanical pain, temporal summation of pain (TSP), conditioned pain modulation (CPM), and cold pain intensity. The sequence of heat and mechanical testing was randomized and counterbalanced, with CPM always administered last to minimize any potential carryover effects. The same researcher performed QST on each participant throughout the study, and all participants received standardized recorded instructions to prevent bias during data collection and enhance the reliability of the results. More details on QST procedures are provided in **Supplemental Material S1.**

#### 2.4.3. Outcome Variable: HTE

Our previous research (blind for review) utilized the multi-trajectory latent class growth analysis (MT-LCGA),^33^ implemented through the SAS *Proc Traj* procedure, to determine distinct latent groups exhibiting similar trajectories based on jointly modeled KOA pain and symptom parameters from baseline to the three-month follow-up post-intervention within the active tDCS group. Detailed methods for MT-LCGA are provided in **Supplemental Material S2**. As a result, a two-group multi-trajectory model was selected to represent two distinct response trajectories (**Table 1 and Figure 2**). The most suitable models for the majority of longitudinal latent classes were characterized by linear slopes. Group 1, labeled “low initial symptoms with significant improvement,” demonstrated significant linear decreases in most symptoms (NRS: β = -0.09, *p* = .037; WOMAC pain: β = -0.01, *p* = .036; and WOMAC stiffness: β = - 0.01, *p* = .001). On the other hand, Group 2, described as “high initial symptoms with minimal improvement,” displayed higher values for all KOA pain and symptoms compared to Group 1. Although Group 2 exhibited a general decline in symptoms over time, these changes were not statistically significant (NRS: β = -0.02, *p* = .599; WOMAC pain: β = -0.01, *p* = .113; WOMAC stiffness: β = -0.004, *p* = .182; WOMAC physical function: β = -0.01, *p* = .518). Group 1 was identified as *high responders*, whereas Group 2 was determined as *low responders*, representing the outcome of the current study. The baseline group characteristics are detailed in **Supplemental Material S3**.

**Table 1.** Two-group multi-trajectory model results.

| | Engagement group | Parameter | $\beta$ (estimate) | Standard error | *t*-value | *p*-value |
| --- | --- | --- | --- | --- | --- | --- |
| Trajectory group (%) | Group 1 |  | 46.91 | 6.55 | 7.17 | <.000 |
|  | Group 2 |  | 53.09 | 6.55 | 8.11 | <.000 |
| NRS pain | Group 1 | linear | -0.09 | 0.04 | -2.08 | **.037** |
|  | Group 2 | linear | -0.02 | 0.04 | -0.53 | .599 |
| WOMAC subscale: pain | Group 1 | linear | -0.01 | 0.01 | -2.10 | **.036** |
|  | Group 2 | linear | -0.01 | 0.01 | -1.59 | .113 |
| WOMAC subscale: stiffness | Group 1 | linear | -0.01 | 0.003 | -3.20 | **.001** |
|  | Group 2 | linear | -0.004 | 0.003 | -1.33 | .182 |
| WOMAC subscale: physical function | Group 1 | linear | -0.03 | 0.02 | -1.27 | .205 |
|  | Group 2 | linear | -0.01 | 0.02 | -0.65 | .518 |

*Note.* Akaike Information Criteria (AIC) = -5320.22, Bayesian Information Criteria (BIC) = -5377.20
Statistically significant *p*-values were written in **bold**.
Group 1 was identified as *high responders*, whereas Group 2 was interpreted as *low responders*.

**Figure 2.**
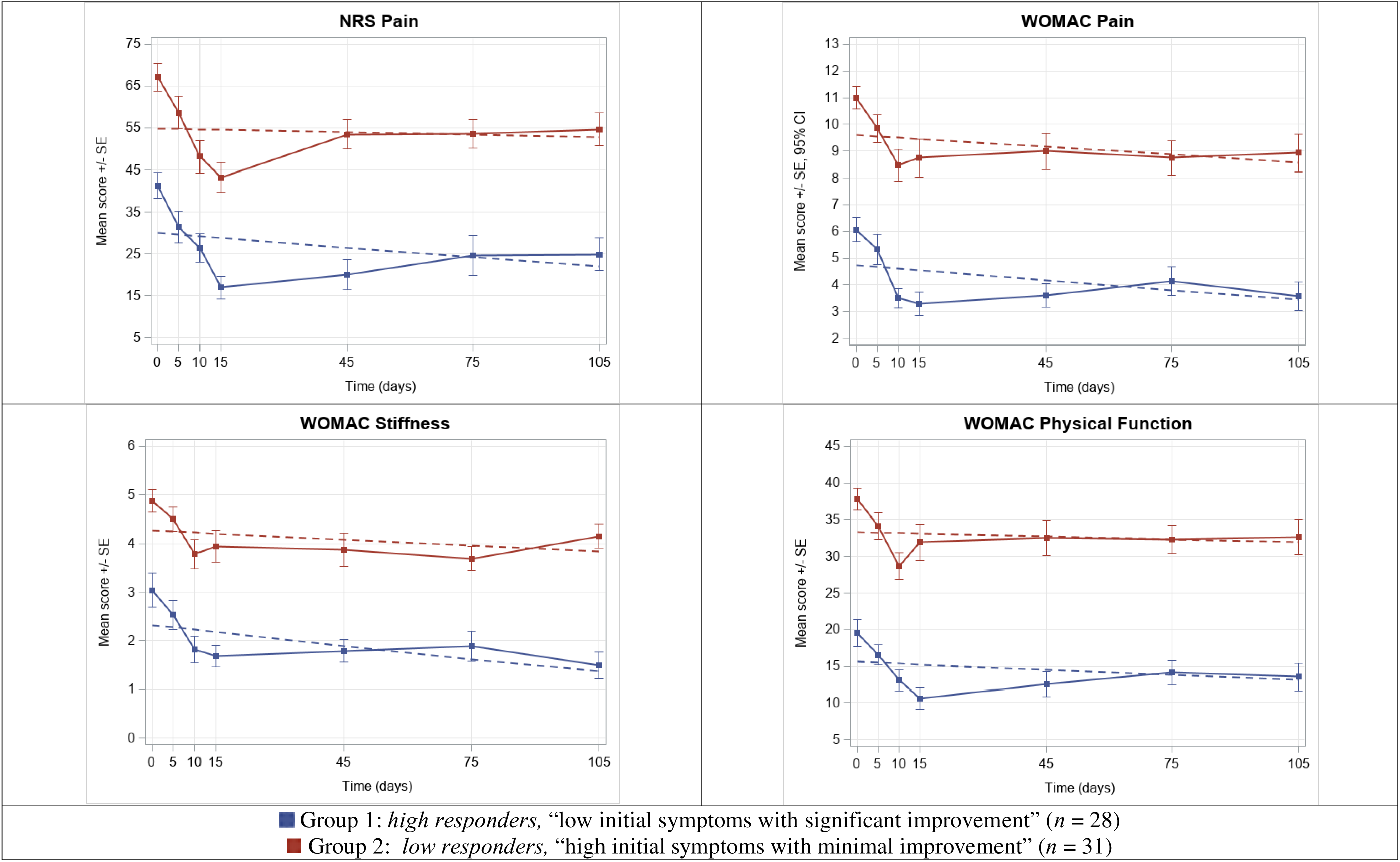
Two-group multi-trajectory model: active tDCS *Note.* The dots represent the mean observed levels, while the solid line represents the expected trajectories. The error bars denote the standard error (SE). *Abbreviation.* NRS, Numeric Rating Scale; tDCS, transcranial direct current stimulation; WOMAC, Western Ontario and McMaster Universities Osteoarthritis Index

### 2.5. Statistical Analysis: ML Approaches

The ML analysis was conducted in Python. Data preprocessing incorporated data imputation and normalization. A MLP classifier with three hidden layers were utilized (**Supplemental Material S4**). Classification performance was evaluated using metrics such as the area under the receiver operating characteristic curve (AUC), accuracy, F-1 score, precision, recall, specificity, and matthews correlation coefficient (MCC) based on four-fold cross-validation.

#### Feature Selection

Feature selection was carried out using f-regression, r-regression, and SHAP (SHapley Additive exPlanations) to identify influential features for classification. f-regression is a univariate linear regression method that evaluates the relationship between each feature and the target variable using F-statistics. r-regression calculates Pearson’s correlation coefficient (*r*-value) to measure the linear association between each feature and the target variable. Both methods are univariate feature selection techniques from Scikit-learn. SHAP is a powerful method for interpreting complex models by breaking down the prediction of each sample into contributions from each feature (Lundberg et al., 2020; Lundberg & Lee, 2017). This analysis aids in understanding how each input feature impacts the model’s predictions, providing insights into the relationships between input features and the output. The SHAP values were visualized through SHAP summary plots, helping interpret the model’s behavior. By comprehending the contribution of each feature, it becomes possible to explain the model’s decisions, identify key drivers of the output, and gain confidence in the model’s predictions. Each method—f-regression, r-regression, and SHAP—ranked features by importance. The top six and bottom six features from each method were compared, and features ranking in the top 25% in at least two methods were selected as the final features for the reliable identification of consistently important features across multiple criteria.

#### The SHAP analysis

A SHAP summary plot was generated using all features, displaying the importance and correlation of each predictive variable. The plot sorted the features on the y-axis sorted by relative importance, with the most influential predictors positioned at the top. In particular, we focused on examining the relevant information for the influential features identified through feature selection in the previous step. Each feature’s contribution was visualized on the plot, with the position of each point (red indicates high feature values) on the x-axis expressing the contribution of individual participants as the SHAP value, with high positive contributions on the far right. The SHAP values represent the importance of each feature’ for a particular prediction, with positive values indicating the feature increased the probability of the predicted outcome (i.e., high responders in this study’s context)—“positive correlations”—and negative values indicating it decreased the probability—“negative correlations.”

## 3. Results

### 3.1. Model Performance Using All Features

The MLP classifier, utilizing all features, achieved an AUC of 0.989 ± 0.005, an accuracy of 0.950 ± 0.012, an F1-score of 0.947 ± 0.012, a precision of 0.938 ± 0.026, a recall of 0.957 ± 0.030, a specificity of 0.944 ± 0.026, and a MCC of 0.901 ± 0.021. These results demonstrated that the MLP classifier accurately predicted trajectories based on the features collected at the baseline visit.

### 3.2. Feature Selection Results

Out of the 21 features analyzed, six features—“CPM at 60 seconds,” “CPM at 30 seconds,” “cold pain intensity,” “PPTh, medial knee,” “PPTh, trapezius,” and “pain catastrophizing”—were identified as the influential features for predicting response trajectories. **Table 2** presents the feature importance rankings determined by f-regression, r-regression, and SHAP for these selected features.

**Table 2.**
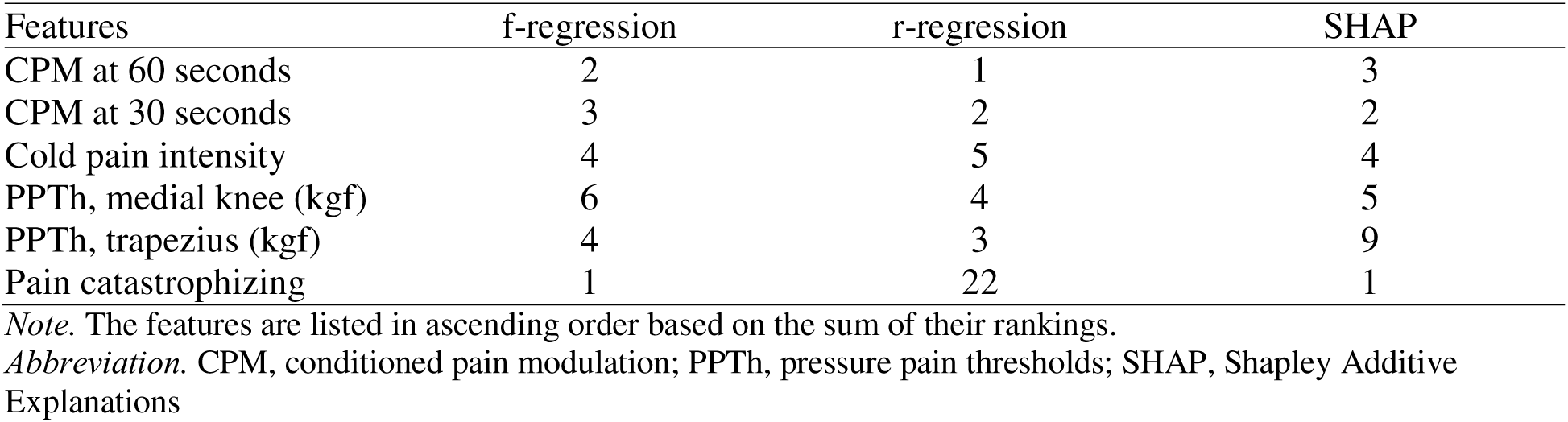
Feature importance rankings.

**Table 3** presents a comparison of MLP classifiers that were trained using different sets of six features. The top and bottom six features represent the most and least important features, respectively, as ranked by f-regression, r-regression, and SHAP. It was observed that MLP classifiers trained with the top six features outperformed those trained with the bottom six features. Furthermore, the MLP classifier utilizing the final selected features demonstrated superior performance compared to the average performance of models using the three feature selection methods in terms of AUC, accuracy, F1-score, and MCC. Specifically, the model with the final selected features achieved an AUC of 0.950, an accuracy of 0.873, an F1-score of 0.861 and a MCC of 0.746, surpassing the average performance metrics (AUC = 0.934, accuracy = 0.857, F1-score = 0.844, MCC = 0.712).

**Table 3.** The MLP classifier performance using six selected features.

| Features | AUC | Accuracy | F1-score | Precision | Recall | Specificity | MCC |
| --- | --- | --- | --- | --- | --- | --- | --- |
| *Top 6 features* |  |  |  |  |  |  |  |
| f-regression | 0.957 | 0.877 | 0.864 | 0.888 | 0.843 | 0.906 | 0.753 |
| r-regression | 0.890 | 0.807 | 0.789 | 0.808 | 0.771 | 0.838 | 0.612 |
| SHAP | 0.954 | 0.887 | 0.879 | 0.879 | 0.879 | 0.894 | 0.772 |
| *Bottom 6 features* |  |  |  |  |  |  |  |
| f-regression | 0.789 | 0.690 | 0.652 | 0.686 | 0.621 | 0.750 | 0.375 |
| r-regression | 0.755 | 0.707 | 0.669 | 0.708 | 0.636 | 0.769 | 0.410 |
| SHAP | 0.899 | 0.780 | 0.776 | 0.742 | 0.814 | 0.750 | 0.564 |
| *Final selected features<sup>a</sup>* | 0.950 | 0.873 | 0.861 | 0.881 | 0.843 | 0.900 | 0.746 |

Note. <sup>a</sup>“CPM at 60 seconds,” “CPM at 30 seconds,” “cold pain intensity,” “PPT<sub>h</sub>, medial knee,” “PPT<sub>h</sub>, trapezius,” and “pain catastrophizing”
Abbreviation. AUC, area under the receiver operating characteristic curve; CPM, conditioned pain modulation; MCC, matthews correlation coefficient; PPT<sub>h</sub>, pressure pain thresholds; SHAP, SHapley Additive Explanations

### 3.3. The SHAP Analysis

Among the final six selected features identified by f-regression, r-regression, and SHAP, “pain catastrophizing” was found to have the largest feature importance value, followed by “CPM at 60 seconds,” “CPM at 30 seconds,” “cold pain intensity,” “PPTh, medial knee,” and “PPTh, trapezius” (**Figure 3).** A positive correlation was observed for features such as “pain catastrophizing” and “cold pain intensity,” indicating that higher values of these features are associated with an increased likelihood of being classified as low responders. In contrast, features such as “CPM at 60 seconds,” “CPM at 30 seconds,” “PPTh, medial knee,” and “PPTh, trapezius” exhibited a negative correlation, suggesting that lower values of these features tend to be associated with an increased likelihood of being classified as low responders.

**Figure 3.**
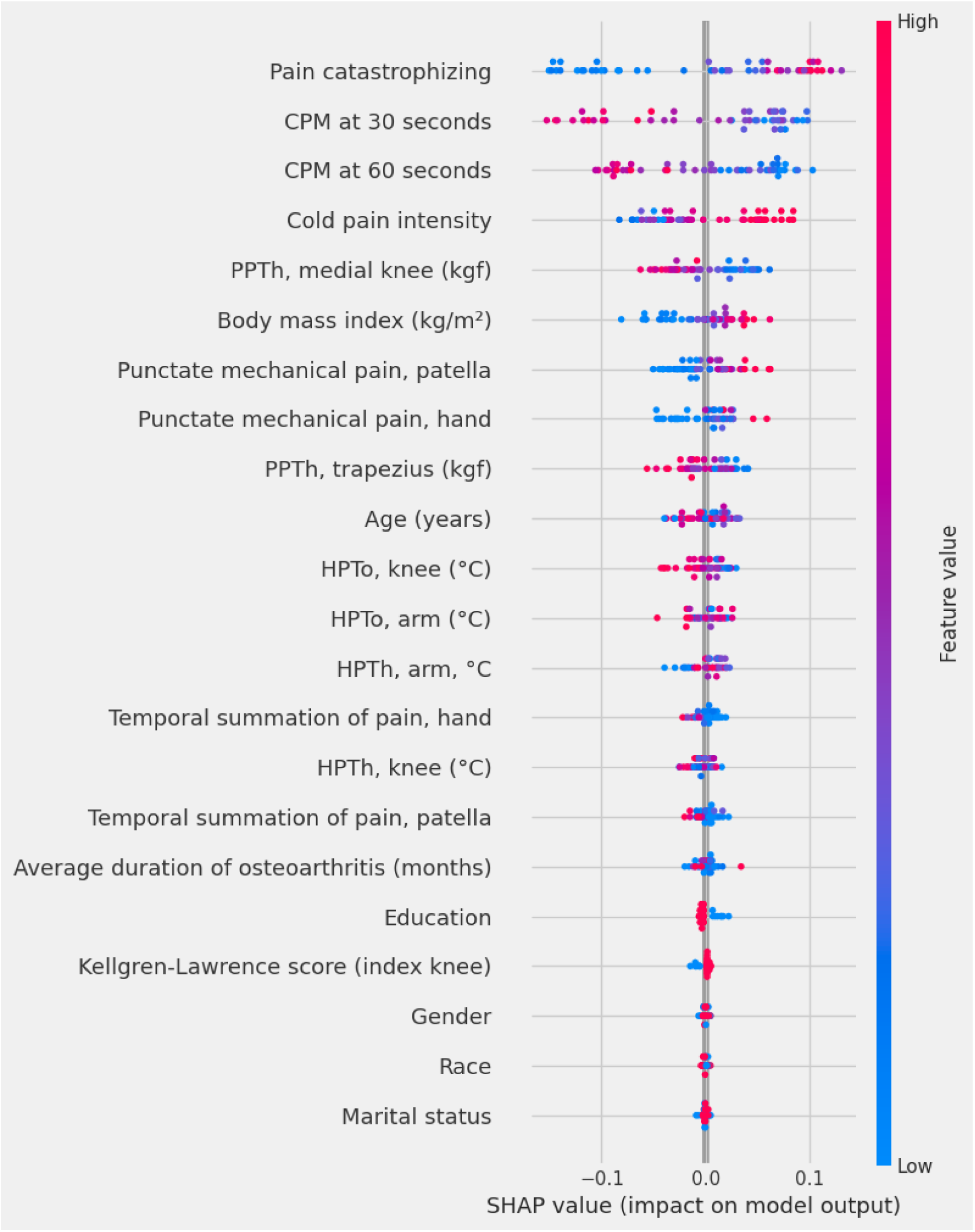
A SHAP summary plot: global interpretation (average feature importance) and local interpretation (SHAP value distribution) of the model. *Abbreviation.* CPM, conditioned pain modulation; PPTh, pressure pain threholds; HPTh, heat pain threshold, HPTo, heat pain tolerance

## 4. Discussion

In this study, the influential baseline features for predicting response trajectories (high responders vs. low responders), selected using f-regression, r-regression, and SHAP, included CPM at 60 seconds, CPM at 30 seconds, cold pain, PPTh at both the medial knee and trapezius, and pain catastrophizing.

These findings underscore the importance of considering these characteristics as crucial moderating factors when developing personalized stimulation protocols. The SHAP analysis provided a visual interpretation of the direction and strength of each feature’s impact on the model’s prediction.

According to the SHAP values, pain catastrophizing, which refers to maladaptive emotional and cognitive response to pain, was identified as the most influential feature in predicting response trajectories. Notably, higher levels of pain catastrophizing were associated with a greater likelihood of being classified as a low responder. This finding corroborates earlier literature indicating that pain catastrophizing is a strong predictor of treatment outcomes for various chronic pain conditions (e.g., low back pain, musculoskeletal pain, and fibromyalgia).^34–36^ Moreover, research suggests that pain treatment programs are more effective when they target and reduce pain catastrophizing.^37^

Given the complexity of chronic pain, in which biological, psychological (emotional and cognitive), and social factors coexist and exacerbate one another, a holistic biopsychosocial approach is theoretically the optimal management strategy.^38^ Based on our findings, it is crucial to consider incorporating pain catastrophizing-oriented therapies alongside tDCS for individuals with high pain catastrophizing. In tDCS research for KOA, M1 is commonly chosen because its stimulation is believed to modulate lateral thalamic hyperactivity, which is associated with the sensory-discriminative aspects of pain processing.^39,40^ In addition to M1, a crucial area involved in pain processing is the dorsolateral prefrontal cortex (DLPFC). The DLPFC is responsible for processing the cognitive and attentional demands associated with painful stimuli and plays a significant role in modulating pain perception as well as pain-related emotional reactions.^41–43^ Furthermore, altered connectivity between the DLPFC and deeper pain-related brain regions,^44^ alongside reductions in grey matter density and DLPFC volume,^45^ is linked to chronic pain. These findings underscore the multifaceted role of the DLPFC in pain perception and its potential as a therapeutic target for modulating the cognitive and emotional dimensions of pain. The left DLPFC is primarily related to cognitive and emotional pain processing. Previous studies have demonstrated that left DLPFC stimulation reduces pain catastrophizing in patients with fibromyalgia.^46,47^ Notably, Gurdiel-Álvarez et al.^48^ have adopted a technique called concurrent dual-site anodal tDCS (a-tDCS), which targets both the DLPFC and M1 region, aiming to harness the potential advantages of motor cortex modulation and cognitive-emotional regulation for a more comprehensive pain relief strategy compared to conventional M1-targeted tDCS methods. Only two studies have investigated the effects of dual-site tDCS over M1 and DLPFC on pain processing: one focused on the endogenous pain inhibitory system among healthy participants,^48^ the other examined pain relief in type-2 diabetes patients with neuropathic pain.^49^ This approach has not yet been explored in individuals with symptomatic KOA and should be examined in future studies. In addition, future research may investigate the possibility of combining tDCS with cognitive behavioral therapy^50^ or meditation.^51^

The results from the SHAP analysis indicate that lower CPM values are associated with a higher likelihood of being classified as low responders. While the role of CPM as a predictive tool has been primarily investigated in pharmacological treatments,^52–54^ its relationship with response to tDCS remains unexplored. Our finding suggests that the endogenous pain inhibitory capacity may have an important role in forecasting treatment outcomes, emphasizing the significance of assessing CPM for treatment selection and prognosis prediction. In addition, high cold pain was linked to a greater likelihood of being classified as low responders. Cold pain intensity is not commonly assessed in KOA; however, some studies have shown higher ratings of pain intensity and unpleasantness at certain temperatures in the high symptomatic KOA group.^55,56^ In this study, increased hypersensitivity to cold pain at the unaffected site may serve as a marker of central sensitization, further supporting the presence of generalized pain hypersensitivity in low responders to tDCS.

The interpretation of SHAP also revealed that lower PPTh at both the medial knee and trapezius are associated with a higher likelihood of being classified as low responders. Lower PPTh at the affected site indicates peripheral sensitization, while lower PPTh at the remote site reflects central sensitization.^32^ These results suggest that both peripheral and central sensitization may predict response to tDCS in KOA patients. Recent studies report a significant prevalence of pain sensitization in KOA patients, estimated at 20%.^57^ Theoretically, patients with low PPTh values are expected to exhibit poor responses to treatment due to sensitized pain mechanisms. In addition, central sensitization, characterized by where various changes in somatosensory input processing mechanisms and overstimulation of central nervous system neurons, can make pain treatment more challenging. For instance, the presence of preoperative central sensitization (e.g., widespread pain sensitization) was related to poor outcomes after total knee replacement.^58^

In light of these findings, it is imperative to consider QST parameters in treatment planning particularly for low responders identified in this study. Individuals with impaired endogenous pain inhibitory capacity or heightened pain sensitivity may benefit from a novel multimodal approach that combines tDCS with other active interventions. An additional therapeutic approach aimed at desensitizing the central nervous system may be beneficial particularly in such individuals. For instance, exercise has been shown to have central analgesic effects by activating descending inhibitory mechanisms^59,60^ and reducing proinflammatory cytokines associated with increased pain sensitization by inducing a cytokine response.^61,62^ Pain neuroscience education can also be integrated into treatment for patients with centrally mediated pain,^63,64^ as it can influence pain perception through descending pain modulation by acting on the dorsal horn of the spinal cord.^65^ Simultaneously, it may be also essential to re-evaluate treatment parameters such as stimulation intensity and duration, session frequency, and intervals between sessions.

Certain limitations should be noted. First, the availability of large, multi-modal datasets poses a substantial constraint on predictive analytics. While the number of participants included in this study is adequate for a neuromodulation randomized controlled trial in KOA, it may be insufficient for drawing robust conclusions in a prediction study. Another limitation is the lack of detailed information in the feature space used to predict response to tDCS. One systematic review supports that depression and anxiety may also mediate the effects of tDCS on chronic pain,^66^ yet our dataset only included pain catastrophizing as a psychological factor. In addition, while demographic, clinical, and QST variables are easily obtainable at a relatively low cost, the absence of neuroanatomical, functional, genetic, or biomarker data in training statistical models may hinder the classification of high responders versus low responders.^67^ Incorporating this data into existing prediction models could further strengthen model performance in the future.^68^ Despite these limitations, our findings offer valuable insights into the HTE of tDCS in KOA populations and highlight the need to prioritize certain approaches to enhance this cost-effective, tolerable, and safe intervention.

## 5. Conclusion

This study identified key predictors of the HTE of tDCS in individuals with symptomatic KOA using ML approaches. These included pain catastrophizing, peripheral and central pain sensitization, and individuals’ endogenous pain-inhibitory capacity—all of which should be scrutinized in future tDCS trials. Pain catastrophizing, in particular, emerged as the strongest predictor mediating the treatment response to tDCS in KOA. This information may offer valuable insights that can be utilized to inform personalized stimulation protocols, potentially improving treatment outcomes.

## Data Availability

All data produced in the present study are available upon reasonable request to the authors.

## Notes

### Competing Interest Statement

The authors have declared no competing interest.

### Clinical Trial

NCT04016272

### Funding Statement

This study was supported by the NIH/NINR Grant R15NR018050.

### Author Declarations

The University of Arizona Institutional Review Board (IRB)

