## Supplemental Materials for "Part 2: Predicting heterogeneity of treatment effects to transcranial direct current stimulation for knee osteoarthritis pain and symptoms"

### **Supplemental Material S1.** QST procedures

**Thermal Testing Procedures.** Contact heat stimuli were delivered using a computer-controlled TSA-II NeuroSensory Analyzer (Medoc Ltd., Ramat Yishai, Israel) to assess HPTh and HPTo at both the index knee and the ipsilateral ventral forearm, employing an ascending method of limits. To prevent sensitization or habituation of cutaneous receptors, the thermode position was rotated among three locations between trials at each body site. Starting at a baseline temperature of 32°C, the thermode temperature increased at a rate of 0.5°C per second until participants pressed a button on a handheld device. Participants were instructed to press the button when they first perceived the heat as painful to measure HPTh and when the heat became intolerable to assess HPTo. Three HPTh trials were conducted at the first test site, followed by three HPTo trials. The same sequence was then repeated at the second test site, with a 5-minute rest period between sites. The average of the three trials was calculated for each participant to determine overall HPTh and HPTo temperatures for analysis.

**Mechanical Testing Procedures.** PPTh was evaluated using a handheld digital pressure algometer (Wagner, Greenwich, Connecticut, USA) to apply blunt mechanical pressure to deep tissues, including muscles and joints. The pressure was increased at a constant rate of 0.3 kgf/cm² per second to determine PPTh at two locations: the medial side of the index knee and the trapezius muscle. The order of testing sites was randomized and counterbalanced. Participants were instructed to indicate the point at which the sensation “first became painful” and at which point the applied pressure was recorded. The average of three trials at each site was calculated to determine the PPTh for that location.

After assessing PPTh, participants underwent testing to evaluate cutaneous mechanical sensitivity to punctate stimuli on both the index patella and back of the ipsilateral hand. A calibrated nylon monofilament delivering a target force of 300 g was applied 10 times at a rate of one contact per second, with participants providing verbal pain intensity ratings on a scale from 0 (no pain sensation) to 100 (the most intense pain imaginable). Pain ratings from the two trials were averaged to punctate mechanical pain at each site. For TSP assessment, participants first rated the pain intensity from a single application of the monofilament. They then rated the maximum pain intensity experienced during a series of 10 contacts administered at a rate of one contact per second. Temporal summation of pain was calculated by subtracting the pain rating for the single stimulus from the rating for the series of 10 stimuli at each site.

**CPM*.*** Ten minutes after assessing thermal or mechanical pain, CPM was evaluated by calculating the change in PPTh during the cold pressor task relative to baseline PPTh. Baseline PPTh measurements were obtained immediately before participants immersed their hands in a cold water bath maintained at 12°C. Thirty seconds after immersion, participants rated their cold pain intensity on a scale from 0 to 100, followed by a second PPTh measurement. Participants were instructed to keep their hands submerged for as long as tolerable for up to a maximum of one minute. Upon hand removal, the final PPTh measurements were recorded. CPM at 30 and 60 seconds was calculated by subtracting baseline PPTh from PPTh at 30 seconds and PPTh at 60 seconds, respectively. The 12°C temperature was selected based on prior studies involving middle-aged and older adults with KOA, as it was found to induce moderate yet tolerable pain in most participants (King et al., 2013). The water was continuously circulated and maintained at a stable temperature using a refrigeration unit (Neslab, Portsmouth, New Hampshire, USA). An increase in PPTh following cold water immersion indicated pain inhibition.

### **Supplemental Material S2.** Multi-trajectory latent class growth analysis (MT-LCGA)

The MT-LCGA was utilized in this study. This approach, implemented through the SAS *Proc Traj* procedure, aimed to determine distinct latent groups exhibiting similar trajectories based on jointly modeled KOA pain and symptom parameters from baseline to the three-month follow-up post-intervention within the active tDCS group. The MT-LCGA approach takes full advantage of the available data, particularly in situations where there is often a strong interrelationship or collinearity among different symptoms (i.e., pain, stiffness, and physical functional disability). Models ranging from one to three latent groups were analyzed and compared. The selection of the optimal number of multi-trajectory groups was based on various fit criteria, including the Akaike Information Criterion (AIC), Bayesian Information Criterion (BIC), homogeneity, latent class separation, interpretability, and group proportion, with a focus on interpretability and group proportion. Lower AIC and BIC values were indicative of better model fit. Each identified group was required to represent at least 10.0% of the sample to allow for meaningful interpretation and further analysis, as very small groups may lack significance or reproducibility. All analyses were conducted using SAS© software, Version 9.4 (SAS Institute Inc., Cary, NC) to ensure accuracy and reliability in the results obtained.

### **Supplemental Material S3.** Baseline group characteristics

|  | *n* (%) or mean ± standard deviation | | |
| --- | --- | --- | --- |
|  | Active tDCS (*n* = 60) | | |
|  | All participants | Group 1  (*n* = 28) | Group 2  (*n* = 32) |
| Age, years | 65.32 ± 8.41 | 66.10 ± 8.57 | 64.63 ± 8.34 |
| Gender |  |  |  |
| Male | 20 (33.3%) | 11 (39.3%) | 9 (28.1%) |
| female | 60 (60.7%) | 17 (58.6%) | 23 (71.9%) |
| Body mass index (kg/m^2^) | 32.67 ± 8.73 | 30.09 ± 6.23 | 34.93 ± 10.00 |
| Race |  |  |  |
| white | 26 (43.3%) | 14 (50.0%) | 12 (37.5%) |
| non-white | 34 (56.7%) | 14(50.0%) | 20 (62.5%) |
| Education |  |  |  |
| high school or less | 13 (21.7%) | 2 (7.1%) | 11 (34.4%) |
| college or more | 47 (78.3%) | 26 (92.9%) | 21 (65.6%) |
| Marital status |  |  |  |
| married/partnered | 38 (63.3%) | 18 (64.2%) | 20 (62.5%) |
| nonmarried/unpartnered | 22 (36.7%) | 10 (35.7%) | 12 (37.5%) |
| Kellgren-Lawrence score (index knee) |  |  |  |
| 0-1 | 9 (15.0%) | 6 (21.4%) | 3 (9.4%) |
| ≥ 2 | 51 (85.0%) | 22 (78.6%) | 29 (90.6%) |
| Average duration of osteoarthritis (months) | 71.35 ± 75.23 | 70.64 ± 74.47 | 71.97 ± 78.24 |
| Pain catastrophizing (PCS) | 15.65 ± 13.99 | 8.21 ± 8.45 | 22.16 ± 14.73 |
| *Quantitative sensory testing* |  |  |  |
| HPTh, knee (°C) | 39.40 ± 3.30 | 39.66 ± 3.53 | 39.17 ± 3.13 |
| HPTh, arm (°C) | 38.37 ± 2.75 | 38.49 ± 3.01 | 38.26 ± 2.54 |
| HPTo, knee (°C) | 44.85 ± 2.74 | 45.54 ± 2.50 | 44.24 ± 2.82 |
| HPTo, arm (°C) | 43.94 ± 3.56 | 44.44 ± 3.21 | 43.51 ± 3.84 |
| PPTh, medial knee (kgf) | 2.40 ± 1.02 | 2.76 ± 0.95 | 2.10 ± 0.99 |
| PPTh, trapezius (kgf) | 2.41 ± 0.93 | 2.80 ± 0.93 | 2.07 ± 0.80 |
| Punctate mechanical pain, patella (average series of 10 in patella punctate mechanical, score: 0-100) | 33.73 ± 33.79 | 23.88 ± 29.25 | 42.36 ± 35.55 |
| Punctate mechanical pain, hand (average series of 10 in patella punctate mechanical, score: 0-100) | 26.55 ± 1.69 | 15.20 ± 19.65 | 29.83 ± 30.03 |
| Temporal summation of pain, patella  (series of 10 - single trial) | 13.39 ± 19.62 | 13.45 ± 18.62 | 13.34 ± 20.76 |
| Temporal summation of pain, hand  (series of 10 - single trial) | 10.52 ± 15.62 | 9.16 ± 3.98 | 11.70 ± 17.49 |
| CPM at 30 seconds  (PPTh at 30 seconds - PPTh at pre-CPM) | 0.44 ± 0.45 | 0.61 ± 0.33 | 0.30 ± 0.50 |
| CPM at 60 seconds  (PPTh at 60 seconds - PPTh at pre-CPM) | 0.60 ± 0.47 | 0.79 ± 0.50 | 0.38 ± 0.34 |
| Cold pain intensity at 30 seconds (score: 0-100) | 77.65 ± 25.06 | 69.11 ± 24.57 | 85.13 ± 23.36 |

*Abbreviation.* CPM, conditioned pain modulation; PCS, pain catastrophizing scale; PPTh, pressure pain threholds; HPTh, heat pain threshold, HPTo, heat pain tolerance; tDCS, transcranial direct current stimulation

### **Supplemental Material S4**

Missing values in the dataset were addressed using the mean imputation method with SimpleImputer, ensuring minimal data loss and maintaining analytical consistency. To enhance model stability and convergence speed, MinMaxScaler was applied to normalize all feature values from 0 to 1. This normalization mitigated scale differences among features, preventing any single feature from disproportionately influencing the model during training.

**Model hyperparameters.** A MLP classifier was trained to predict response trajectories using demographic, clinical, and QST data collected at baseline. The hyperparameters were fine-tuned to optimize model performance, with the MLP architecture consisting of three hidden layers with 256, 128, and 64 neurons, respectively. To introduce non-linearity, the ReLU activation function was applied, and the Adam optimizer was used for weight updates. The learning rate was set to 0.0001 to ensure stable convergence, with a maximum of 10 iterations. The training process spanned 20 epochs to maximize the model’s generalization performance, with a batch size of 45 ensuring the entire dataset was used in a single training iteration.

**Performance optimization.** The model was optimized to enhance its predictive performance on the entire dataset (*n* = 60). Due to the limited dataset size (*n* = 60), four-fold cross-validation and random noise augmentation were employed to ensure performance enhancement (**Table S1**). Additionally, hyperparameter tuning was first performed on one of the four folds to obtain the optimal model performance. The best hyperparameters were then applied uniformly across all folds. In four-fold cross-validation, the 15 predicted outputs from each fold were aggregated, and the final performance evaluation was conducted based on all 60 samples.

To further enhance model performance, random noise was introduced, which was integrated into the training data while eliminating all other sources of randomness, with the exception of noise generation. To determine the optimal standard deviation (SD) for the random noise, performance was evaluated across various SD values. It was observed that when the SD exceeded 1, both training and validation performance deteriorated. Values below 1 were tested with 10 different noise scales (0, 0.001, 0.0025, 0.005, 0.01, 0.025, 0.05, 0.1, 0.25, 0.5), and the average performance was computed over five different seeds.

Among the six performance metrics, AUC, accuracy, and F1-score—commonly considered for model selection—were prioritized. As a result, optimal performance was achieved with a noise scale of 0.005 (**Table S2**).

### **Table S1.** Data sampling composition

| Fold/Group | Active tDCS | |
| --- | --- | --- |
|  | High responders | Low responders |
| 1 fold – train | 23 | 22 |
| 1 fold – test | 10 | 5 |
| 2 fold – train | 25 | 20 |
| 2 fold – test | 8 | 7 |
| 3 fold – train | 25 | 20 |
| 3 fold – test | 8 | 7 |
| 4 fold – train | 26 | 19 |
| 4 fold - test | 8 | 7 |

*Note.* The dataset consisted of 60 samples in the active tDCS group. A four-fold cross-validation approach was utilized, with each fold containing 45 training samples and 15 test samples from each group. The distribution of samples per class in the active tDCS is presented in the Table**.** Given the constraints of medical datasets, a four-fold cross-validation strategy was implemented to ensure all samples were utilized in both training and validation processes. Missing values in the dataset were addressed using the mean imputation method with SimpleImputer, minimizing data loss and maintaining analytical consistency. To enhance model stability and convergence speed, MinMaxScaler was applied to normalize all feature values within the range of 0 to 1. This normalization mitigated scale differences among features, preventing any single feature from disproportionately influencing the model during training. To enhance the generalization performance of the model and prevent overfitting during training, random noise was introduced to the training data, following a standard normal distribution.

### **Table S2.** Model performance results by noise scale

| **Noise scale** | **AUC** | **Accuracy** | **F1 score** | **Precision** | **Recall** | **Specificity** | **MCC** |
| --- | --- | --- | --- | --- | --- | --- | --- |
| 0 | 0.990 | 0.950 | 0.947 | 0.944 | 0.950 | 0.950 | 0.900 |
| 0.001 | 0.988 | 0.950 | 0.947 | 0.938 | 0.957 | 0.944 | 0.900 |
| 0.0025 | 0.988 | 0.947 | 0.943 | 0.938 | 0.950 | 0.944 | 0.894 |
| **0.005** | **0.989** | **0.950** | **0.947** | **0.938** | **0.957** | **0.944** | **0.901** |
| 0.01 | 0.989 | 0.947 | 0.943 | 0.938 | 0.950 | 0.944 | 0.894 |
| 0.025 | 0.986 | 0.943 | 0.940 | 0.931 | 0.950 | 0.938 | 0.887 |
| 0.05 | 0.987 | 0.943 | 0.940 | 0.931 | 0.950 | 0.938 | 0.887 |
| 0.1 | 0.988 | 0.947 | 0.944 | 0.938 | 0.950 | 0.944 | 0.894 |
| 0.25 | 0.990 | 0.947 | 0.945 | 0.914 | 0.979 | 0.919 | 0.896 |
| 0.5 | 0.973 | 0.910 | 0.905 | 0.890 | 0.921 | 0.900 | 0.820 |
